# Risk of Group 2 Pulmonary Hypertension in Newly Diagnosed Heart Failure: An EHR-Based Cohort Analysis

**DOI:** 10.64898/2025.12.03.25341274

**Authors:** Zeshui Yu, Yuqing Chen, Manling Zhang, Ning Feng, Lirong Wang, Yu Chen, Flora Sam

## Abstract

**Background:** Group 2 pulmonary hypertension (PH) is associated with poor clinical outcomes. Comprehensive epidemiological data on the incidence of Group 2 PH following heart failure (HF) with preserved versus reduced ejection fraction (HFpEF *vs*. HFrEF) are limited.

**Objectives:** To investigate the differential associations of HFpEF *vs.* HFrEF with incident Group 2 PH.

**Methods:** 17,212 newly diagnosed HF patients were examined from University of Pittsburgh Medical Center between 10/01/2015 and 03/31/2021. The cumulative incidence of PH was estimated using the Nelson-Aalen method, and associations with clinical risk factors were assessed using Cox proportional hazards models. Men and women were analyzed separately, given established sex differences in HF subtypes.

**Results:** Mean age was 71.7±13.0 years, 46.3% were women, and 1,636 incident cases of PH were identified up to 4.5-years of follow-up. The cumulative incidence of PH was initially higher in HFrEF but was surpassed by HFpEF after 2 years. The cumulative incidence (95% CI) was 33.51% (32.36%–34.65%) in HFpEF and 30.87% (29.04%–32.65%) in HFrEF. The incidence was higher in women 36.25% (34.84%–37.62%) than in men 29.69% (28.33%–31.02%). HFpEF in women was associated with a 23% higher risk of incident PH *vs.* HFrEF (HR: 1.23; 95% CI: 1.01–1.50). This differential risk by HF subtype was not seen in men.

**Conclusion:** This study highlights the significant burden of the development of PH in HF with HFpEF showing higher risk, especially in women. These findings suggest that HF subtype and sex may influence PH development through distinct pathways.

**Condensed abstract:** Group 2 pulmonary hypertension (PH) worsens heart failure (HF) outcomes, but its incidence by HF subtype is unclear. In 17,212 UPMC HF patients, PH was initially more common in HFrEF but surpassed by HFpEF after two years. Women had higher PH incidence, with HFpEF increasing PH risk by 23% in women but not in men. These findings highlight the need for PH screening in all HF patients, especially those with HFpEF and women. Integrating PH risk stratification into HF care may support earlier intervention, guide treatment decisions, and improve long-term outcomes in this growing, high-risk population.

## INTRODUCTION

Pulmonary hypertension (PH) is a pathophysiological condition characterized by elevated pressures in the pulmonary vasculature.^1^ It is a common and severe complication in patients with heart failure (HF), and its presence is almost always associated with worse symptoms, concurrent serious comorbidities, frequent hospitalizations, and increased mortality.^2, 3^ Although resting right heart catheterization (RHC) is the gold standard for diagnosing and classifying PH, defined as a resting mean pulmonary artery pressure (mPAP) >20 mmHg and a pulmonary capillary wedge pressure (PCWP) >15 mmHg^4^ non-invasive methods such as echocardiography are increasingly used as screening tools. However, their utility remains limited due to variability in how mPAP is estimated across populations with differing heart failure (HF) etiologies and severities.^5^ Prior studies report a prevalence of PH in patients with HF with preserved ejection fraction (HFpEF) as ∼36-83%, and that of HF with reduced ejection fraction (HFrEF) as ∼40-75%.^6^

Secondary PH is a progressive condition and a poor prognostic indicator of worsening HF.^7, 8^ It is typically classified as Group 2 PH or post-capillary PH associated with left heart disease (PH-LHD).^5^ This classification accounts for ∼65-80% of all PH cases, due to the widespread prevalence of left heart disease.^2^ The primary mechanism involves a sustained elevation of left atrial pressure (LAP) from impaired left ventricular (LV) function, leading to chronically elevated pulmonary filling pressures and resultant congestion within the pulmonary veins and capillaries.^8^ Despite its clinical relevance, large-scale real-world clinical data on the incidence of Group 2 PH in newly diagnosed HF patients are lacking. ^7, 8^ Prior studies largely focused on the prevalence of PH in small studies with limited sample sizes in patients with established HF and its association with biomarkers, rather than that of clinical characteristics.^9–11^

The clinical characteristics and presentation of Group 2 PH in HF vary significantly depending on whether it is associated with HFpEF or HFrEF.^12, 13^ To date, the differential association between HFpEF versus HFrEF and the risk of incident Group 2 PH has not been investigated in real-world HF patients. Currently, there are also no established therapies specifically targeting PH in HFpEF, and treatment is directed primarily towards addressing symptoms and underlying comorbidities.^14^ Conversely, those with decompensated or end-stage HFrEF who develop resting PH and high filling pressures may respond in a limited fashion to acceleration of guideline-directed therapy for HFrEF or medical therapy that attempts to normalize pulmonary pressure while maintaining systemic pressures and cardiac output.^15, 16^ Additionally, the interplay between comorbidities and PH in the context of HF is considered complex and involves multiple factors.^2^ Interestingly, several observational studies had suggested that 55-65% of HFpEF patients were women.^17, 18^ Thus, these findings pose the question of whether sex contributes to the risk or progression of PH. Taken together, these gaps underscore the need to identify potential risk factors to further elucidate the pathophysiology of Group 2 PH in HF patients, particularly in relation to HF subtypes and sex.

We therefore leveraged a large, real-world cohort of newly diagnosed HF patients from electronic health records (EHR) with longitudinal follow-up. Our study had three main objectives : (i.) estimate the incidence of Group 2 PH in this cohort; (ii.) assess the differential associations of HFpEF versus HFrEF with the development of Group 2 PH; and (iii.) identify key clinical risk factors associated with incident Group 2 PH. Additionally, we conducted sex-stratified analyses to investigate potential differences in risk factors and disease progression between men and women.

## METHODS

### Study design and patient eligibility

The University of Pittsburgh Medical Center (UPMC) has one of the largest EHR systems in the United States and serves as a resource for translational research. This EHR provides real-world patient data derived from several healthcare settings. Notably, it includes data sources from UPMC Presbyterian Hospital, a high-acuity, tertiary care facility recognized for its clinical care in cardiovascular and pulmonary medicine. This study received approval from the Institutional Review Board (IRB) at the University of Pittsburgh (STUDY19020153, approved on March 13th, 2019). We conducted a retrospective cohort study that included all patients (n=40,404) who were older than 20 years old and initially diagnosed with HF based on the documented ICD codes (**Supplemental Table 1**) between October 1^st^, 2015, and March 31^st^, 2021, in the UPMC healthcare system. Complete patient information, including demographics, diagnosis, laboratory tests, etc., was extracted from the UPMC EHR system. The date of the initial HF diagnosis in the UMPC health care system served as the **index date** for the study, and patients were then followed until one of the following came first: (i.) the occurrence of incident Group 2 PH, (ii.) death, (iii.) the last encounter, (iv.) the end of a 4.5-year follow-up and (v.) March 31^st^ , 2021 (**Figure 1**). Group 2 PH was identified based on the first recorded occurrence of specific ICD-10 codes in Supplemental Table 1. If a patient had a diagnosis code of I27.23 or I27.24, the diagnosis date was defined as the earliest instance of either of these codes or any of the following related codes: I27.2, I27.20, I27.22, or I27.29. For patients without I27.23 or I27.24, the diagnosis was based on the earliest occurrence of any of the latter four codes, as shown in **Supplemental Table 1.**

**Figure 1.**
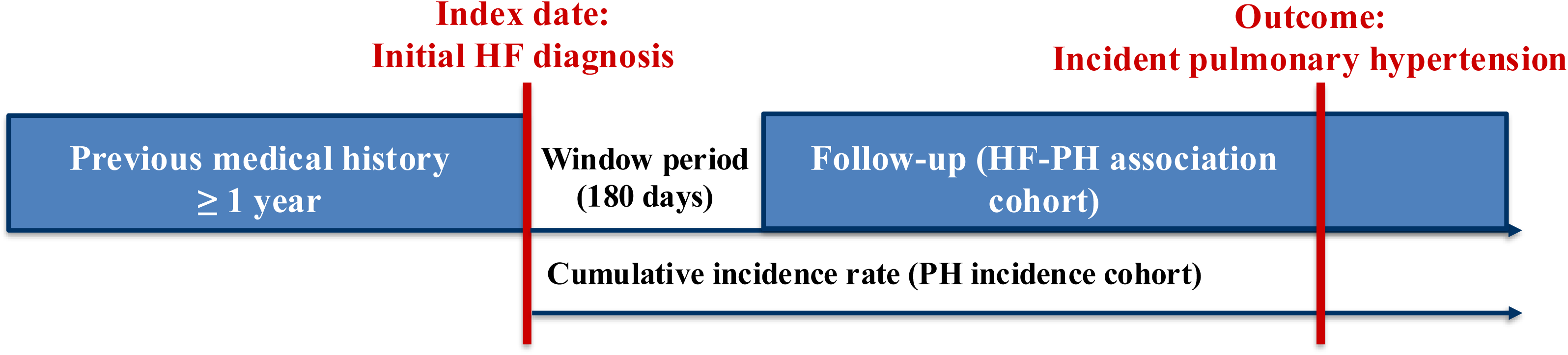

For the analysis, several inclusion and exclusion criteria were established to ensure reliable and meaningful results (**Figure 2**). (**i.**) To be classified as having newly diagnosed HF, patients were required to have at least one year of medical history prior to the index date and, crucially, have no HF diagnosis codes at any point in their available medical record before the index date. (n=31,646). (**ii.**) Patients had their baseline body mass index (BMI) determined as the closest value recorded within the window of 180 days before *and* up to 7 days after the index date of the initial HF diagnosis (n=29,232). (**iii.**) Patients with any ICD codes indicating primary PH or pulmonary arterial hypertension (PAH) also known as Group 1 PH were excluded from the analysis as this is a different disease compared to Group 2 PH. Exclusion was based on the presence of these codes at any time or in any record within the EHR (n=28,156). Group 1 PH is a rare disease with precapillary involvement and/or idiopathic/heritable genetic mutations with multiple effective treatments and a different pathogenesis from Group 2 PH where there is no recommended approved guideline-directed therapy for the latter.^19^ (**iv.**) Patients with ICD codes indicating Group 3 or Group 4 PH (I27.23 or I27.24) and without additional coding indicating Group 2 PH during the follow-up were excluded (n = 28,019). These codes were identified during the follow-up after the HF index date. (**v.**) Patients were required to have a documented left ventricle ejection fraction (LVEF) within 365 days before and after the index date (n=22,280). The availability of LVEF measurement helped ensure that the HF diagnosis was clinically validated. (**vi**.) Patients with a pre-existing PH diagnosis (on or before the HF index date) were excluded. (n=17,970). (**vii**.) Patients were required to have follow-up, defined as at least one encounter after the index date (n=17,212)

**Figure 2.**
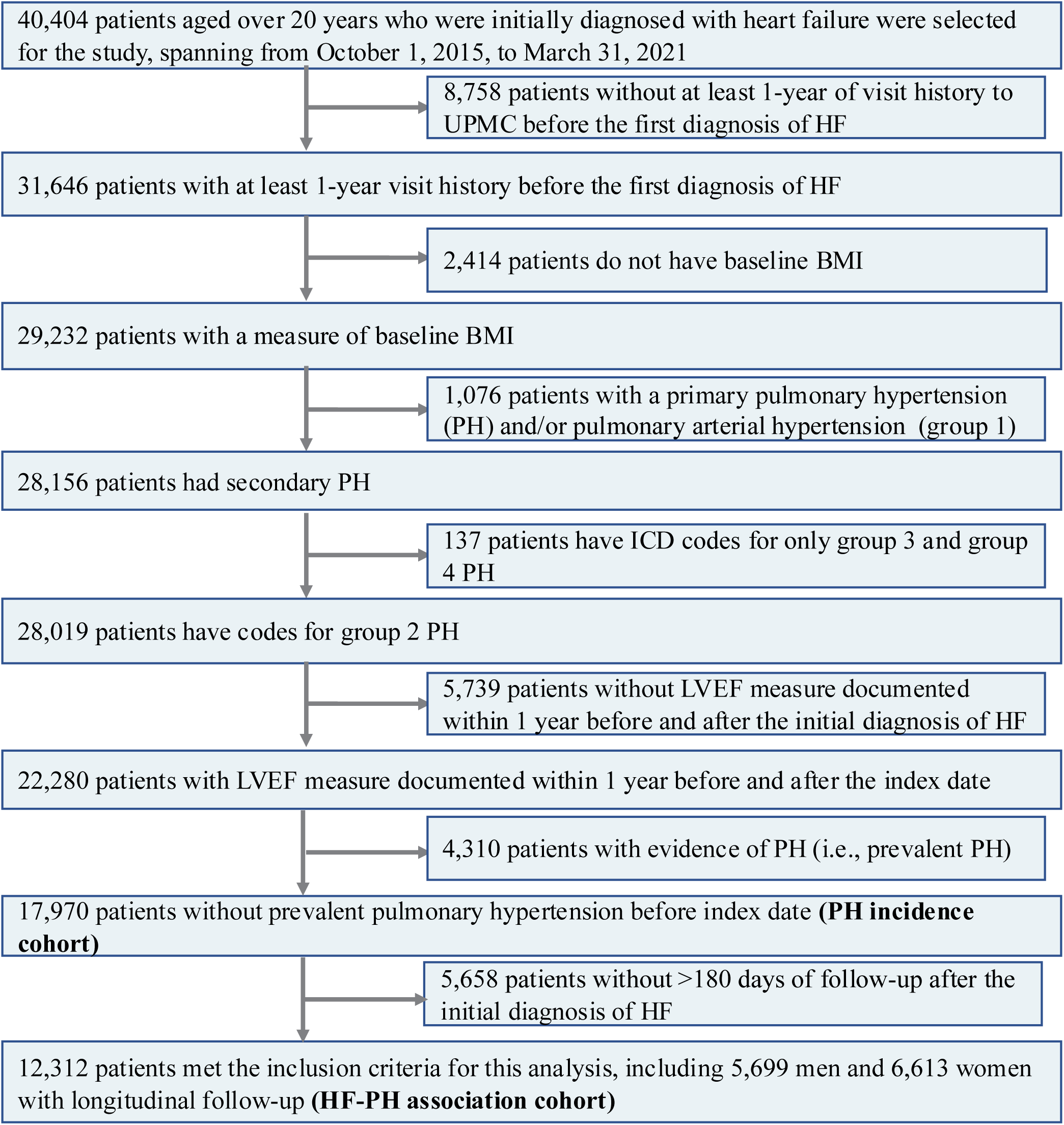
Inclusion flowchart.

We selected this cohort of 17,212 individuals to calculate the incidence of Group 2 PH diagnosed clinically following the first diagnosis of HF (**PH incidence cohort, Figure 1**). To explore the association between potential risk factors and incident Group 2 PH, we established a 180-day window period following the initial diagnosis of HF to include patients diagnosed with Group 2 PH or who had continued follow-up during this time (n=12,312) (**HF-PH association cohort, Figure 1**). This time frame was implemented to address frequent diagnostic delays of prevalent PH because RHC is not routinely performed in HF patients, and the development of PH is a progressive process post-HF.^8, 20, 21^ Additionally, our analysis below shows that prevalent PH is more likely to be diagnosed shortly after an HFrEF diagnosis compared to HFpEF, potentially due to the ease of HFrEF diagnosis based on a depressed LV ejection fraction, symptoms of congestion leading to early medical attention, and undergoing tests for PH.^20^ The study design and inclusion flowchart of the patient selection are shown in **Figure 2**.

### Selection and Identification of Potential Risk Factors

Several components of the EHR were used to extract relevant variables for this analysis. These included patient demographics such as age at the index date, gender, race, and comorbidities. Race is categorized as white, black, or other. The other race comprises individuals identifying as Asian, Pacific Islander, Middle Eastern, mixed Native American, or mixed race. A comprehensive list of comorbidities was selected as the potential risk factors for evaluation based on published literature and clinical expertise from cardiologists. These include anemia, atrial fibrillation (Afib), chronic kidney disease (CKD), chronic obstructive pulmonary disease, coronary arterial disease, hyperlipidemia, hypertension, hyperthyroidism, nonalcoholic fatty liver disease or nonalcoholic steatohepatitis, obstructive sleep apnea, peripheral arterial disease, type 2 diabetes (T2D), and venous thromboembolism. The presence of comorbidities was identified using ICD-9 and ICD-10 codes in the diagnosis file before or at the initial date of HF diagnosis.

A full list of codes has been derived from previous literature and is shown in **Supplemental Table 1**. The subtype of HF is classified based on the LVEF values obtained from the most recent echocardiogram performed within one year before or after the index date, where HFrEF was defined as an LVEF ≤ 40%, and HFpEF was defined by an LVEF > 40%. While HFpEF is theoretically defined as a LVEF > 50%, our analysis included patients with LVEF between 40 and 50% with HFpEF. Patients with such an LVEF are known as HF with midrange ejection fraction (HFmrEF). HFmrEF patients were included as they share many clinical characteristics as well as common comorbidities, such as hypertension and Afib with HFpEF.^22^ Additionally, outcomes for HFmrEF patients are more aligned with those for HFpEF than HFrEF.^23^

### Statistical analysis

Descriptive statistical analysis was performed to evaluate the baseline demographics and characteristics of patients by the development of Group 2 PH and by HF subtype among both incidence and HF-PH association cohorts separately. Continuous variables were summarized using the mean ± standard deviation (SD), while categorical variables were presented as count and percentage (%). Two-sample t-test and chi-square test were applied for continuous variables and categorical variables, respectively, to compare the differences between the groups. We estimated the cumulative incidence and 95% confidence interval (CI). We used the Nelson-Aalen estimator to estimate the cumulative incidence of Group 2 PH following HF and 95% CI during the follow-up in the entire incidence cohort, by sex and by HF subtype.

For the time-to-event analysis in HF-PH association cohorts, follow-up began 180 days after the index date to account for potential diagnostic delays of preexisting PH and the increased likelihood of PH being diagnosed in patients with HFrEF, given the presence of symptoms such as volume overload and pulmonary congestion.^8, 20, 21^ The proportion of patients remaining at risk of developing incident Group 2 PH was estimated by Kaplan-Meier models in the entire HF-PH association cohorts and stratified by sex in a sex-stratified analysis. To identify potential risk factors associated with incident Group 2 PH, univariate Cox proportional hazards regression models were conducted in the sex-pooled analysis. Variables with a p-value < 0.1 were included in the multivariable Cox models. Subsequently, sex-stratified multivariable Cox models were used to assess the independent associations of these risk factors with incident Group 2 PH, adjusting for age, race, BMI, and other relevant covariates. A two-sided *p-*value < 0.05 was deemed statistically significant. All analyses were conducted using Python 3.10 and SAS 9.4.

## RESULTS

### Baseline Characteristics of Study Cohort

After implementing the inclusion criteria, we identified a cohort of patients for the PH incidence analysis. Descriptive statistics for the demographics and baseline characteristics of this cohort, categorized by incident PH development and by HF subtype, are shown in **Supplemental Table 2**. After the application of the 180-day window period, 12,312 patients were enrolled in the HF-PH association cohort, with an average follow-up of 1.65 years beyond the window. Demographics and baseline characteristics are shown in **Table 1**. Within this cohort, 9,472 (76.9%) patients were classified as HFpEF, and 2,840 (23.1%) as HFrEF. The cohort’s mean age was 71.7 years (SD: 13.0), comprising 5,699 (46.3%) female patients and 6,613 (53.7%) male patients. Over a median follow-up of 1.93 years (SD: 1.18), 1,636 (13.4%) patients developed PH. Patients who developed incident Group 2 PH were older, female, and more likely to be Black compared to those who did not develop Group 2 PH.

**Table 1.** Demographics and Characteristics of the HF-PH Association Cohort.

|  |  | Entire HF-PH association cohort | No incident PH | Incident PH | p-Value | HFpEF | HFrEF | p-Value |
| --- | --- | --- | --- | --- | --- | --- | --- | --- |
| n |  | 12312 | 10676 | 1636 |  | 9472 | 2840 |  |
| Age, mean (SD) |  | 71.7 (13.0) | 71.3 (12.9) | 74.0 (12.9) | <0.005 | 72.4 (12.7) | 69.4 (13.5) | <0.005 |
| Gender, n (%) | Female | 5699 (46.3) | 4851 (45.4) | 848 (51.8) | <0.005 | 4681 (49.4) | 1018 (35.8) | <0.005 |
|  | Male | 6613 (53.7) | 5825 (54.6) | 788 (48.2) |  | 4791 (50.6) | 1822 (64.2) |  |
| Race, n (%) | White | 11128 (90.4) | 9690 (90.8) | 1438 (87.9) | <0.005 | 8630 (91.1) | 2498 (88.0) | <0.005 |
|  | Black | 1062 (8.6) | 886 (8.3) | 176 (10.8) |  | 748 (7.9) | 314 (11.1) |  |
|  | Other | 122 (1.0) | 100 (0.9) | 22 (1.3) |  | 94 (1.0) | 28 (1.0) |  |
| Body Mass Index mean (SD) |  | 31.2 (7.8) | 31.2 (7.8) | 31.2 (7.8) | 0.85 | 31.6 (7.9) | 29.8 (7.1) | <0.005 |
| Heart Failure subtype, n (%) | HFpEF | 9472 (76.9) | 8178 (76.6) | 1294 (79.1) | 0.03 | N/A | N/A |  |
|  | HFrEF | 2840 (23.1) | 2498 (23.4) | 342 (20.9) |  |  |  |  |
| Anemia, n (%) |  | 4920 (40.0) | 4168 (39.0) | 752 (46.0) | <0.005 | 4032 (42.6) | 888 (31.3) | <0.005 |
| Atrial fibrillation, n (%) |  | 5001 (40.6) | 4213 (39.5) | 788 (48.2) | <0.005 | 3886 (41.0) | 1115 (39.3) | 0.097 |
| Coronary artery disease, n (%) |  | 8337 (67.7) | 7253 (67.9) | 1084 (66.3) | 0.19 | 6294 (66.4) | 2043 (71.9) | <0.005 |
| Chronic kidney disease, n (%) |  | 4050 (32.9) | 3399 (31.8) | 651 (39.8) | <0.005 | 3238 (34.2) | 812 (28.6) | <0.005 |
| Chronic obstructive pulmonary disease, n (%) |  | 5001 (40.6) | 4311 (40.4) | 690 (42.2) | 0.18 | 4014 (42.4) | 987 (34.8) | <0.005 |
| Hyperlipidemia, n (%) |  | 10462 (85.0) | 9085 (85.1) | 1377 (84.2) | 0.35 | 8165 (86.2) | 2297 (80.9) | <0.005 |
| Hypertension, n (%) |  | 11630 (94.5) | 10054 (94.2) | 1576 (96.3) | <0.005 | 9040 (95.4) | 2590 (91.2) | <0.005 |
| Hypothyroidism, n (%) |  | 3359 (27.3) | 2861 (26.8) | 498 (30.4) | <0.005 | 2739 (28.9) | 620 (21.8) | <0.005 |
| Nonalcoholic fatty liver disease or nonalcoholic steatohepatitis, n (%) |  | 1039 (8.4) | 917 (8.6) | 122 (7.5) | 0.14 | 854 (9.0) | 185 (6.5) | <0.005 |
| Obstructive sleep apnea, n (%) |  | 3148 (25.6) | 2756 (25.8) | 392 (24.0) | 0.12 | 2597 (27.4) | 551 (19.4) | <0.005 |
| Peripheral arterial disease, n (%) |  | 4000 (32.5) | 3437 (32.2) | 563 (34.4) | 0.08 | 3235 (34.2) | 765 (26.9) | <0.005 |
| Type 2 diabetes, n (%) |  | 5817 (47.2) | 4973 (46.6) | 844 (51.6) | <0.005 | 4580 (48.4) | 1237 (43.6) | <0.005 |
| Venous thromboembolism, n (%) |  | 1328 (10.8) | 1150 (10.8) | 178 (10.9) | 0.93 | 1083 (11.4) | 245 (8.6) | <0.005 |

Values are presented as mean ± standard deviation or number (%).
PH: pulmonary hypertension
N/A: not applicable

Furthermore, a significantly higher prevalence of anemia, Afib, CKD, hypertension, hyperthyroidism, and T2D was observed in patients who developed Group 2 PH. Demographics and characteristics of patients were categorized by the subtype of HF, and significant differences were noted in all potential risk factors. Patients with HFpEF exhibited a higher prevalence of metabolic conditions and obesity, while those with HFrEF were more likely to have concurrent cardiovascular diseases.

### Cumulative Incidence of PH Stratified by Gender and HF Type

**Table 2** reports the cumulative incidence of PH after the initial diagnosis of HF over the 4.5-year follow-up period in the incidence cohort. The incidence rates are presented overall and according to specific time periods of interest, stratified by sex and HF subtypes. At 1 year following HF, the cumulative incidence of PH was higher in patients with HFrEF (14.21%, [13.19%, 15.21%]) compared to those with HFpEF (12.43%, [11.88%, 12.97%]). By the end of the second year, the rates were comparable for both subtypes (HFrEF: 19.64%, [18.46%, 20.81%] vs HFpEF: 19.62%, [18.95%, 20.28%]), but by 4.5 years, HFpEF patients had a higher cumulative incidence of PH (33.51%, [32.36%, 34.65%]) compared to HFrEF patients (30.87%, [29.04%, 32.65%]). Importantly, women consistently exhibited a higher rate of PH than men across all observed time periods, with rates at 4.5 years of 36.25% ([34.84%, 37.62%]) for women and 29.69% ([28.33%, 31.02%]) for men. Kaplan-Meier survival curves of incident Group 2 PH in the PH-incidence group were shown in **Figure S1**. A crossover survival curve between HFpEF and HFrEF was observed. To further elucidate these relationships, we evaluated the incidence rate of PH among HF patients, with further stratification by gender within each HF subtype. The analysis revealed that the incidence of PH was persistently higher in women with HFpEF compared to men. In contrast, men and women with HFrEF exhibited similar rates. The detailed results from this analysis are presented **in Supplemental Table S3.**

**Table 2.** Cumulative Incidence % and 95% CIs of Group 2 Pulmonary Hypertension Following Heart Failure.

| **Time from HF** | **All** | **Male** | **Female** | **HFrEF** | **HFpEF** |
| --- | --- | --- | --- | --- | --- |
| 30 days | 3.08 (2.82–3.33) | 2.81 (2.48–3.14) | 3.40 (3.00–3.79) | 3.82 (3.24–4.39) | 2.85 (2.57–3.13) |
| 90 days | 5.80 (5.46–6.14) | 5.27 (4.82–5.71) | 6.42 (5.89–6.94) | 6.79 (6.05–7.53) | 5.49 (5.11–5.87) |
| 180 days | 8.60 (8.19–9.00) | 7.81 (7.27–8.34) | 9.52 (8.90–10.13) | 10.07 (9.19–10.95) | 8.14 (7.69–8.59) |
| 1 year | 12.85 (12.37–13.33) | 11.67 (11.03–12.31) | 14.21 (13.48–14.93) | 14.21 (13.19–15.21) | 12.43 (11.88–12.97) |
| 2 years | 19.63 (19.05–20.21) | 17.95 (17.16–18.74) | 21.53 (20.66–22.38) | 19.64 (18.46–20.81) | 19.62 (18.95–20.28) |
| 3 years | 24.78 (24.11–25.45) | 22.92 (21.99–23.84) | 26.86 (25.89–27.82) | 24.29 (22.92–25.63) | 24.92 (24.15–25.69) |
| 4 years | 29.85 (29.03–30.67) | 27.17 (26.04–28.29) | 32.76 (31.57–33.92) | 29.24 (27.58–30.87) | 30.04 (29.09–30.98) |
| 4.5 years | 32.86 (31.88–33.83) | 29.69 (28.33–31.02) | 36.25 (34.84–37.62) | 30.87 (29.04–32.65) | 33.51 (32.36–34.65) |

The cumulative incidence rate of group 2 pulmonary hypertension in the entire PH incidence cohort after the index date. HF: heart failure; HFrEF: heart failure with reduced ejection fraction; HFpEF: heart failure with preserved ejection fraction

### Differential Associations of HFpEF versus HFrEF on Incident Group 2 PH After HF diagnosis and Risk Factors for Men and Women

First, we conducted a univariate analysis to assess the association between individual potential risk factors and the incidence of Group 2 PH within the sex-pooled HF-PH association cohort. In univariate analysis, HFpEF was associated with a 16% higher risk of incident Group 2 PH (HR: 1.16 [1.03-1.30]), as shown in **Table 3**. There was a significant impact of gender on the risk of developing PH, where male is associated with an 18% lower risk of PH compared to female **(Table 3).** In **Figure 3A (cumulative incidence in Figure S2A),** the survival probability with incident Group 2 PH in the HF-PH cohort is shown with p value from the log-rank test, along with the number of patients at risk at each time interval.

**Figure 3.**
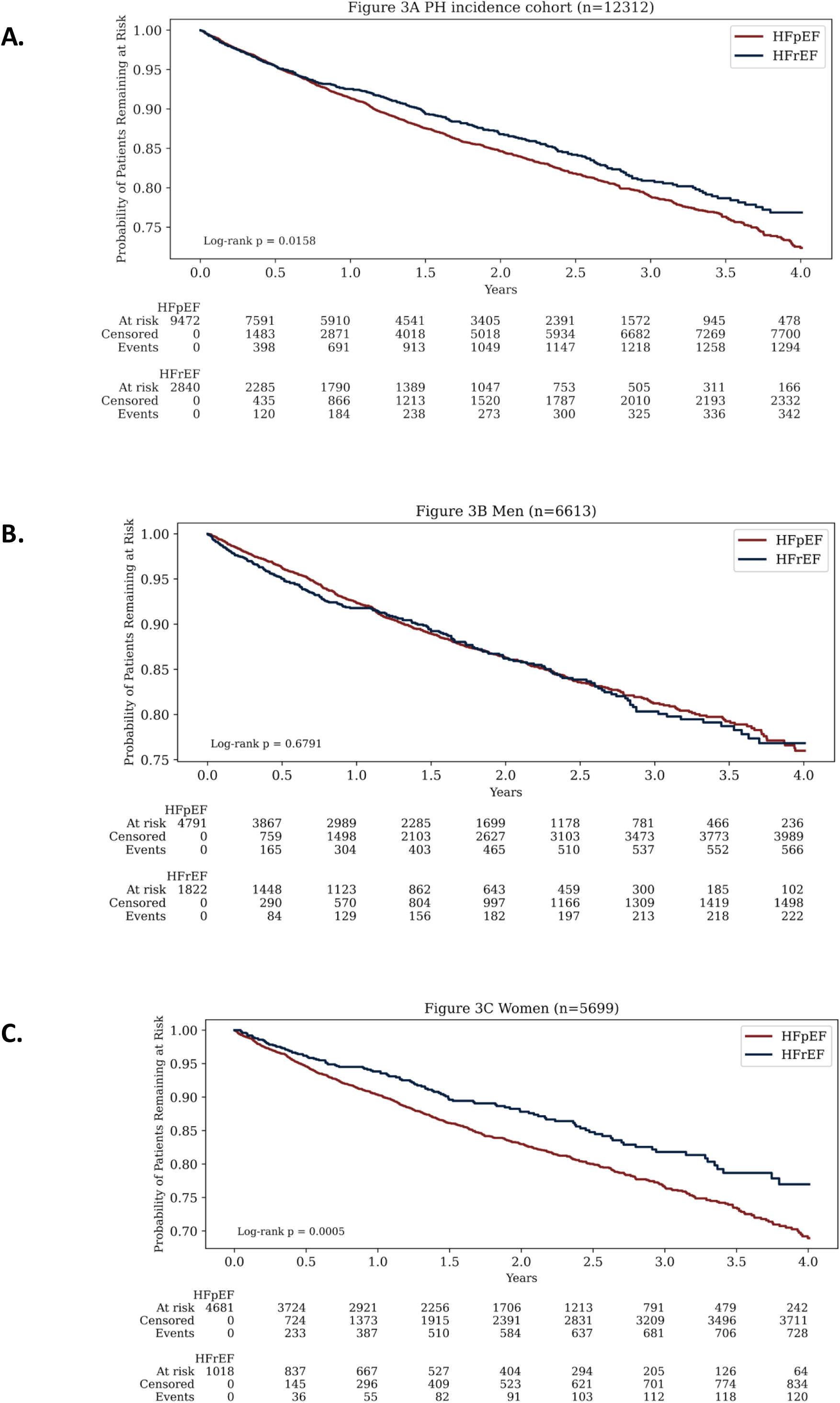
Kaplan-Meier Curve. Estimating the Cumulative probability of Remaining Free from incident Group 2 Pulmonary Hypertension in HF-PH association cohort and in Men and Women **Central Illustration.** Risk of Group 2 Pulmonary Hypertension in Heart Failure: An Electronic Health Record-Based Cohort Analysis

**Table 3.** Associations between Risk Factors and Incident Pulmonary Hypertension in Patients with Heart Failure.

|  |  | Sex-pooled univariate analysis |  | Sex-Stratified multivariate analysis |  |  |  |
| --- | --- | --- | --- | --- | --- | --- | --- |
|  |  | HF-PH Association Cohort (n=12,312) |  | Men (n=6,613) |  | Women (n=5,699) |  |
|  |  | Hazard Ratio | P value | Hazard Ratio | P value | Hazard Ratio | P value |
| Age |  | 1.02 (1.02-1.02) | <0.005 | 1.02 (1.01 - 1.03) | <0.005 | 1.02 (1.01 - 1.02) | <0.005 |
| Race | White | Reference | Reference | Reference | Reference | Reference | Reference |
|  | Black | 1.19 (1.02-1.39) | 0.03 | 1.47 (1.16 - 1.88) | <0.005 | 1.35 (1.08 - 1.69) | 0.01 |
|  | Other | 1.52 (1.00-2.32) | 0.05□ | 1.83 (1.03 - 3.24) | 0.04 | 1.42 (0.76 - 2.65) | 0.27 |
| Gender | Female | Reference | Reference | N/A | N/A | N/A | N/A |
|  | Male | 0.82 (0.74-0.90) | <0.005□ | N/A | N/A | N/A | N/A |
| Body mass index |  | 1.01 (0.99-1.00) | 0.28□ | 1.00 (0.99 - 1.01) | 0.96 | 1.01 (1.00 - 1.02) | 0.20 |
| HFpEF |  | 1.16 (1.03-1.30) | 0.02 | 0.90 (0.77 - 1.05) | 0.19 | 1.23 (1.01 - 1.50) | 0.04 |
| Anemia |  | 1.27 (1.16-1.40) | <0.005 | 1.09 (0.94 - 1.26) | 0.27 | 1.04 (0.91 - 1.20) | 0.57 |
| Atrial fibrillation |  | 1.46 (1.32-1.61) | <0.005 | 1.31 (1.13 - 1.51) | <0.005 | 1.44 (1.25 - 1.66) | <0.005 |
| Chronic kidney disease |  | 1.49 (1.35-1.64) | <0.005 | 1.35 (1.17 - 1.57) | <0.005 | 1.31 (1.13 - 1.52) | <0.005 |
| Chronic obstructive pulmonary disease |  | 1.13 (1.02-1.24) | 0.02 | 1.08 (0.93 - 1.25) | 0.31 | 1.12 (0.98 - 1.29) | 0.10 |
| Coronary artery disease |  | 0.98 (0.89-1.09) | 0.71 | N/A | N/A | N/A | N/A |
| Hyperlipidemia |  | 0.95 (0.83-1.09) | 0.48 | N/A | N/A | N/A | N/A |
| Hypertension |  | 1.78 (1.38-2.31) | <0.005 | 1.12 (0.78 - 1.61) | 0.55 | 1.43 (0.97 - 2.11) | 0.07 |
| Hypothyroidism |  | 1.17 (1.05-1.29) | <0.005 | 0.92 (0.76 - 1.10) | 0.36 | 1.07 (0.94 - 1.23) | 0.31 |
| Nonalcoholic fatty liver disease or nonalcoholic steatohepatitis |  | 0.94 (0.78-1.13) | 0.50 | N/A | N/A | N/A | N/A |
| Obstructive sleep apnea |  | 0.90 (0.80-1.01) | 0.07 | 1.08 (0.93 - 1.26) | 0.28 | 0.97 (0.81 - 1.16) | 0.71 |
| Peripheral arterial disease |  | 1.19 (1.08-1.32) | <0.005 | 0.88 (0.74 - 1.05) | 0.16 | 1.04 (0.90 - 1.21) | 0.61 |
| Type 2 diabetes |  | 1.23 (1.12-1.35) | <0.005 | 1.13 (0.97 - 1.31) | 0.11 | 1.20 (1.04 - 1.38) | 0.01 |
| Venous thromboembolism |  | 0.98 (0.84-1.14) | 0.78 | N/A | N/A | N/A | N/A |

Risk of incident pulmonary hypertension in univariate and multivariable analyses stratified by sex in PH-HF association cohort. Multivariable models were adjusted for age, race, sex, and other relevant clinical covariates identified as potential risk factors (p-value<0.1 in univariate analysis).
PH: pulmonary hypertension; HF: heart failure; HFpEF: heart failure with preserved ejection fraction; N/A: not applicable

Sex-stratified analyses were performed to better understand sex differences in the association between HF subtypes and incidence Group 2 PH. Kaplan-Meier curves of the probability of patients remaining free from incident Group 2 PH in men and women are presented in **Figure 3B and Figure 3C**, respectively **(cumulative incidence in Figure S2B and Figure S2C)**. The risk factors with a p-value less than 0.1 were selected as covariates for multivariate analysis. In men, after adjusting for covariates, there was no significant difference in the risk of developing incident Group 2 PH post-HF between patients with HFpEF and those with HFrEF (HR: 0.90 [0.77-1.05]) (**Table 3**). Each SD increase in age was associated with a 2% increased risk of incident PH (HR: 1.02 [1.01 - 1.03]). Additionally, in men, Black patients and other races exhibited a higher risk of incident PH (HR: 1.47 [1.16-1.88] and 1.83 [1.03-3.24], respectively). Finally, the presence of Afib and CKD in men at baseline was also associated with an increased risk of developing incident Group 2 PH after HF (HR: 1.31 [1.13-1.51] and 1.35 [1.17-1.57], respectively).

In women, the HF subtype plays a significant role in the progression to subsequent Group 2 HF. Multivariable analyses showed that the risk of developing Group 2 PH post-HF was 23% higher in those with HFpEF compared to those diagnosed with HFrEF (HR: 1.23 [1.01 - 1.50]). Each SD increase in age correlated with a 2% increased risk of incident PH (HR: 1.02 [1.01 - 1.03]). Like men, women with Afib (HR: 1.44 [1.24-1.66]) and CKD (HR: 1.31 [1.13-1.52]) had a significantly higher risk of Group 2 PH after HF. Additionally, a higher risk of developing PH was observed in women with T2D concurrent at the index date (HR: 1.20 [1.04-1.39]) that was not seen in men.

## DISCUSSION

A detailed evaluation of the incidence of Group 2 PH was conducted in this large cohort of patients with newly diagnosed HF, and the associations between HF subtypes and risk factors were examined. First, we observed a crossover in the cumulative incidence of Group 2 PH between HFpEF and HFrEF. Patients with HFrEF exhibited higher incidence rates of PH during the first two years of follow-up, but thereafter, the incidence rates for HFpEF exceeded those of HFrEF until the end of the study at 4.5 years. Second, we identified an important sex difference, with HFpEF significantly increasing the risk of incident PH specifically in women, using a window period designed to ensure participants were without PH at the initial diagnosis of HF. Multiple risk factors have been observed to be associated with the development of PH, including CKD and Afib. Our study provides important epidemiological and clinical insights into the progression of PH and highlights the differential impact of heart failure subtypes, particularly in relation to sex-specific risk patterns.

Group 2 PH is increasingly recognized as a serious complication of HF and, unlike that of primary PH or PAH, there are no direct disease-modifying therapies currently available. Additionally, managing these patients in the clinical setting is extremely challenging as the presence of Group 2 PH in HF typically indicates progression to advanced HF, characterized by declining cardiac function with chronic increases in LAP.^20^ Compared to general HF patients, Group 2 PH patients are older with a higher burden of comorbidities, as shown in **Table 1**. To the best of our knowledge, our study represents the first longitudinal study to quantify the incidence of Group 2 PH among newly diagnosed HF patients, considering PH following diagnoses of either HFpEF or HFrEF subtypes. Findings from prior studies vary due to inconsistent definitions of PH—wide ranging from diagnosis codes to echocardiographic assessments to RHC and differing inclusion or exclusion criteria for primary PH or PAH, often using various cutoff points. Many of these prior studies focused on patients with established HF, typically identified through hospitalization or referral for further evaluation. Importantly, Group 2 PH develops as a complication following the onset of HF. To this end, we retrospectively focused on patients newly diagnosed with HF without prevalent PH and used ICD-9 and ICD-10 codes to identify the occurrence of incident PH across the overall HF population, HF subtypes, and by sex to provide real-world clinical evidence. We found the cumulative incidence of PH following HF over 4.5 years of follow-up was 32.86%. These observations provide important data for estimating the future burden of PH in patients with HF in clinical settings, which could aid in healthcare planning and resource allocation. This is especially important given the rising prevalence, increasing hospitalization rate, and the associated high expenses of managing HF.^20,24^

One new observation was an early higher cumulative incidence of Group 2 PH in HFrEF patients compared to HFpEF in the first two years following the initial HF diagnosis. However, as time progressed, the cumulative incidence in HFpEF exceeded that of HFrEF until the end of the study. This crossover in cumulative incidence may be attributed to the natural history of Group 2 PH after HF diagnosis. PH is more easily identified and diagnosed shortly after a clinical presentation of HFrEF due to common signs and symptoms, such as congestion and edema, leading to increased LAP, ease of HFrEF diagnosis with non-invasive imaging, and the availability of numerous guideline-directed therapies for HFrEF.^20^ Conversely, HFpEF is often diagnosed later and so PH becomes apparent later, such as during exertion as a result of steep increases in LAP and cardiac output, thus making PH a timely diagnosis challenging.^25^ This observed crossover in cumulative incidence provides valuable real-world evidence for the difference in diagnosis and progression of PH among HF subtypes.

The initial sex-pool univariate analysis importantly shows that women compared to men have a significantly higher risk of Group 2 PH after HF. The sex-stratified analysis revealed a significantly heterogeneous association between HF subtypes and the incidence of PH, demonstrating that HFpEF was associated with a 23% higher incidence rate of Group 2 PH in women. However, no significant differential impact of the HF subtype was seen in men. It has been suggested that the natural history of PH arising as a complication after HFpEF versus HFrEF may differ. A recent study found that the combination of PH with HFpEF patients was significantly associated with increased in-hospital mortality but not with HFrEF patients.^26^ Few epidemiological studies have explored the differential impact of HF subtypes on the development of Group 2 PH following HF, primarily due to the lack of longitudinal data with imaging such as standardized echocardiogram results. Importantly, the lower risk of PH associated with women with HFrEF may be attributed to the foundational pillars of guideline-directed medical therapies such as ACEIs/ARBs/ARNIs, β-blockers, MRAs, and SGLT2s, which can favorably affect pulmonary hemodynamics by increasing cardiac output and reducing filling pressures, potentially leading to normalization of PAP.^27^ Similarly patients with Group 2 PH in HFrEF who have high resting PH and high filling pressures once decompensated or with progression to end-stage HF, may respond better to medical therapy, than those with HFpEF, resulting in normalization of filling pressures and PA pressures with preserved cardiac output. It has been suggested that the prevalence of Group2 PH also differs depending on the subtype of HF, being highest in HFpEF where multi-comorbidity is common.^28^

Several key risk factors were identified for the development of Group 2 PH with newly diagnosed HF patients, including older age, Black race, Afib, and CKD in both men and women. Our findings are in line with others where the prevalence of PH is increased in patients with CKD as the severity of the kidney disease progresses.^29^ Additionally, a 4-year retrospective analysis of 225 patients demonstrated that Afib in PH was associated with elevated right atrial (RA) pressure and RA dilation, which supports our findings.^30^ Elevated atrial pressure is a pathophysiologic hallmark of HFpEF. Chronically elevated LAP leads to LA enlargement, which further impairs LA function and increase pulmonary pressures and increases the propensity to Afib.^31^ Additional research is necessary to examine the mechanisms underlying these associations and to prevent such high-risk individuals with these comorbidities from developing Group 2 PH. Interestingly, in our study patients with well-described risk factors for PH, such as chronic obstructive pulmonary disease and venous thromboembolism, did not show a significant association with an increased risk of PH.^32^ However the propensity to either Group 3 PH ( i.e., PH due to lung disease) or Group 4 PH ( i.e., PH due to chronic thromboembolic disease) was not investigated in our study. It is possible that early detection and treatment of HF subtypes and the concurrent comorbidities/risk factors could potentially delay or prevent the progression to PH, highlighting the complex interplay of multiple factors determining the likelihood of eventually developing PH in HF patients. The findings in our study also indicate that T2D is a risk factor for incident PH in women but not in men, suggesting potential sex-based differences in the underlying pathobiological mechanisms.

Our study has several significant findings. *First,* it encompasses a large and racially diverse patient cohort, identified by clinical diagnostic ICD-9 and ICD-10 codes with HF subtypes confirmed by echocardiogram, leveraging the EHR system at UPMC. *Second,* this study targets newly diagnosed HF patients. This focus is critical as Group 2 PH development is typically a progressive process following the initial cardiac diagnosis and the cardiac remodeling that occurs in HFrEF and HFpEF.^33^ Currently, very limited real-world clinical evidence is available for the longitudinal follow up from HF diagnosis to PH detection, as most research has focused on prevalent PH in established HF with no longitudinal data. Our approach aims to address this gap by investigating the disease trajectory and its clinical contributors. *Third,* we implemented a unique 180-day window period in our study, designed to ensure patients were free from prevalent PH. This methodological approach is critical for minimizing bias, considering the common delay in HF diagnosis until acute HF-related symptoms occur.^34, 35^ At the acute and initial index HF presentation, prevalent PH may already be developing. Additionally, given the progressive nature of PH following HF and the diagnostic delays due to the lack of routine RHC in HF patients, the likelihood of prevalent PH is high if detected soon after initial HF diagnosis.^20^ Furthermore, prevalent PH is more likely to be diagnosed in HFrEF than in HFpEF during the initial or subsequent visits due to persistent symptoms of congestion despite medical therapy up titration or concurrent hypotension. ^12^

The current study should be interpreted with several potential limitations: (i.) A primary study limitation is the reliance on ICD codes for PH identification, without confirmation by right heart catheterization. This likely introduced misclassification, as some cases were coded as unspecified or other secondary PH, and may have led to over- or underestimation of the true incidence. Despite our institution’s expertise in PH, future studies should combine ICD codes with hemodynamic data to ensure more accurate case identification. (ii.) We are unable to evaluate the severity of both HF and PH due to the absence of hemodynamic parameters. As such pre-capillary and post-capillary PH in Group 2 PH occurs in more advanced and chronic cases but we are unable to discriminate them from post-capillary PH alone (iii.). Patients with prevalent PH at initial HF diagnosis were excluded from analysis, potentially omitting a unique phenotype of Group 2 PH that coincides with HF onset. (iv.) Given our observational study design, we cannot establish causal relationships between identified risk factors and PH development

## Conclusion

In conclusion, in this large, longitudinal, and real-world cohort of patients newly diagnosed with HF, we demonstrated that the cumulative incidence of Group 2 PH reached 32.86% over 4.5 years. Notably, HFpEF conferred a greater risk of developing Group 2 PH in women, a pattern not observed in men, suggesting underlying sex-specific pathobiological mechanisms. The findings in this study highlight the substantial burden of Group 2 PH among HF patients and underscore the need for heightened vigilance in monitoring high-risk individuals. Recognizing these differential impacts across HF subtypes and sex-related disparities may facilitate earlier identification of at-risk patients and guide the development of sex-specific interventions to mitigate risk and improve long-term outcomes.

### PERSPECTIVES

#### Clinical Competency in Medical Knowledge and Patient Care

Group 2 PH due to heart failure is underdiagnosed but once the diagnosis is made, it is associated with increased morbidity and mortality. Although there are presently no disease modifying therapies, early diagnosis, particularly in HFpEF, is important as the transition from isolated postcapillary Group 2 PH to combined postcapillary and precapillary PH denotes advanced disease and increased mortality.

#### Translational Outlook implications

Increased comorbidities in HFpEF increase the propensity to Group 2 PH particular in women, the elderly and those with T2D. Identifying such subtypes within HFpEF- in addition to disease recognition -could aid in the development of novel therapies for Group 2 PH especially due to HFpEF.

## Data Availability

All data produced in the present study are available upon reasonable request to the authors

## Acknowledgements

We thank colleagues from the Health Record Research Request (R3) team of the University of Pittsburgh and the University of Pittsburgh Center for Research Computing and Data for assistance with data access and computational resources.

## ABBREVIATIONS

PH: Pulmonary hypertension
RHC: Right heart catheterization
mPAP: Mean pulmonary artery pressure
PCWP: Pulmonary wedge pressure
PH-LHD: Pulmonary hypertension associated with left heart disease
PAH: Pulmonary arterial hypertension
LAP: Left atrial pressure
LV: Left ventricular
RA: Right atrial
EHR: electronic health records

## SUPPLEMENTAL MATERIAL DOCUMENT

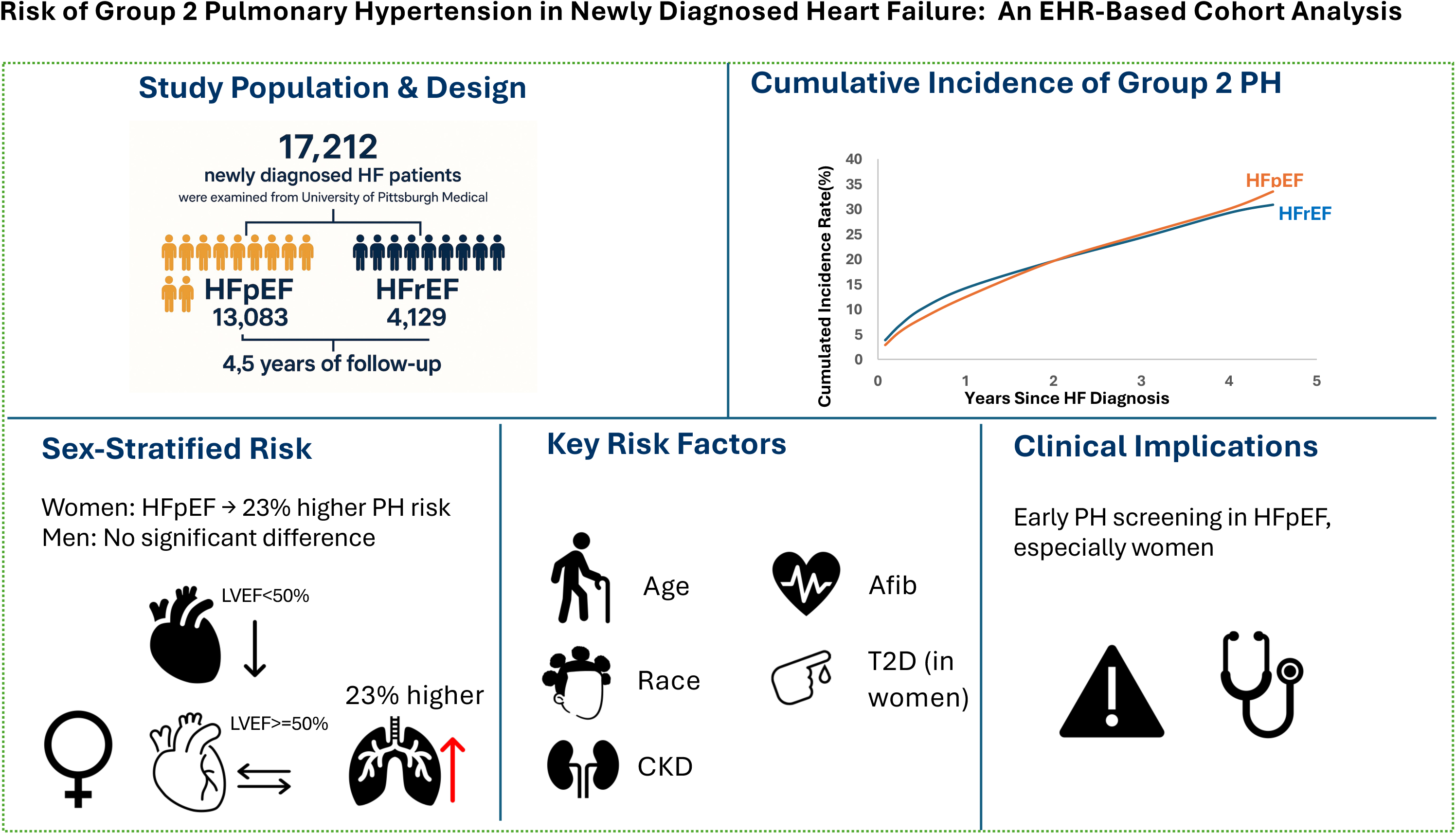

**Supplemental Figure 1.**
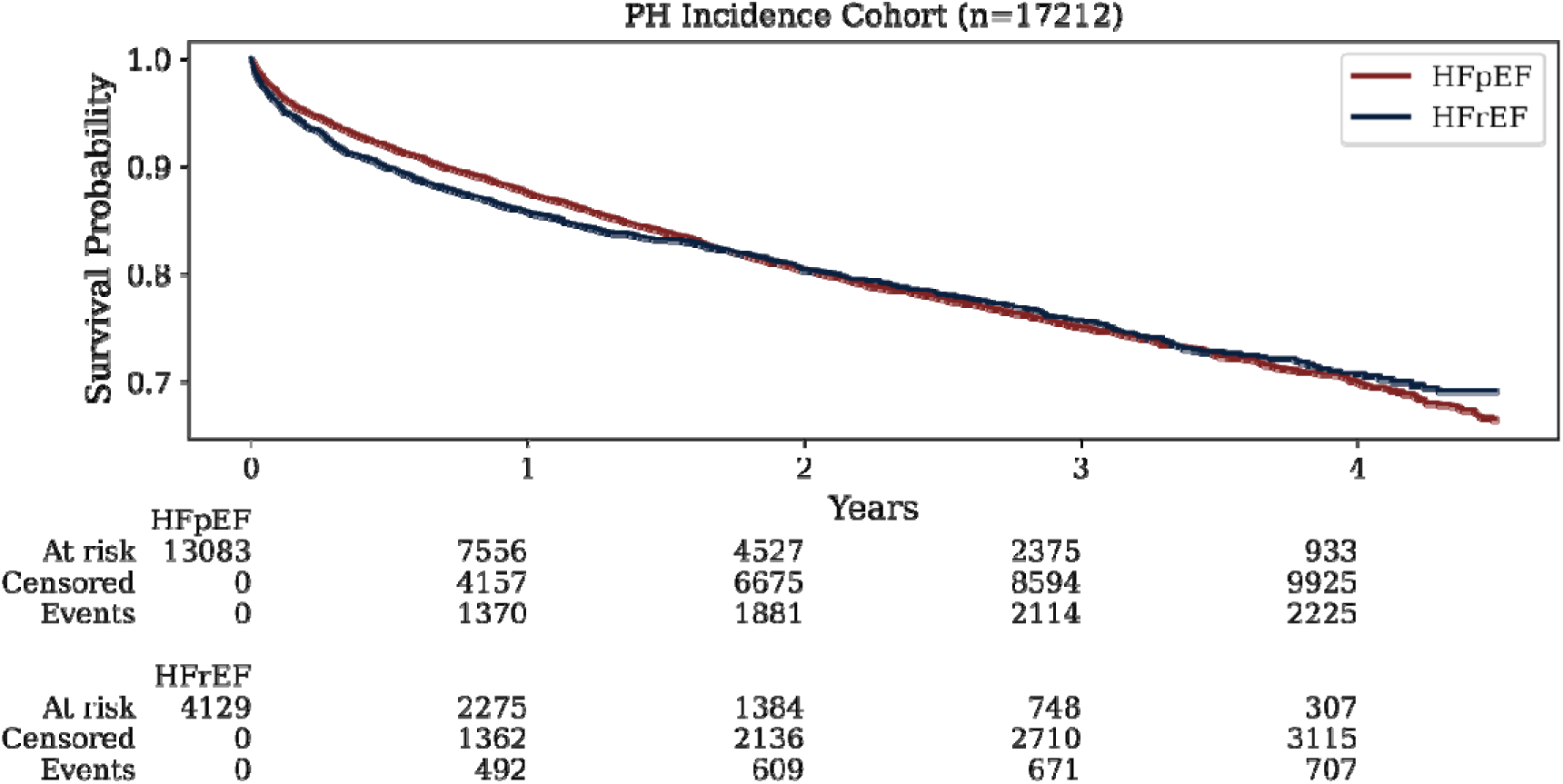
Kaplan-Meier survival curves of incident group 2 PH in the PH-incidence group. Kaplan-Meier survival curve estimating the cumulative probability of remaining free from incident group 2 pulmonary hypertension in the PH-incidence cohort. PH: pulmonary hypertension; HFpEF: heart failure with preserved ejection fraction; HFrEF: heart failure with reduced ejection fraction.

**Supplemental Figure 2.**
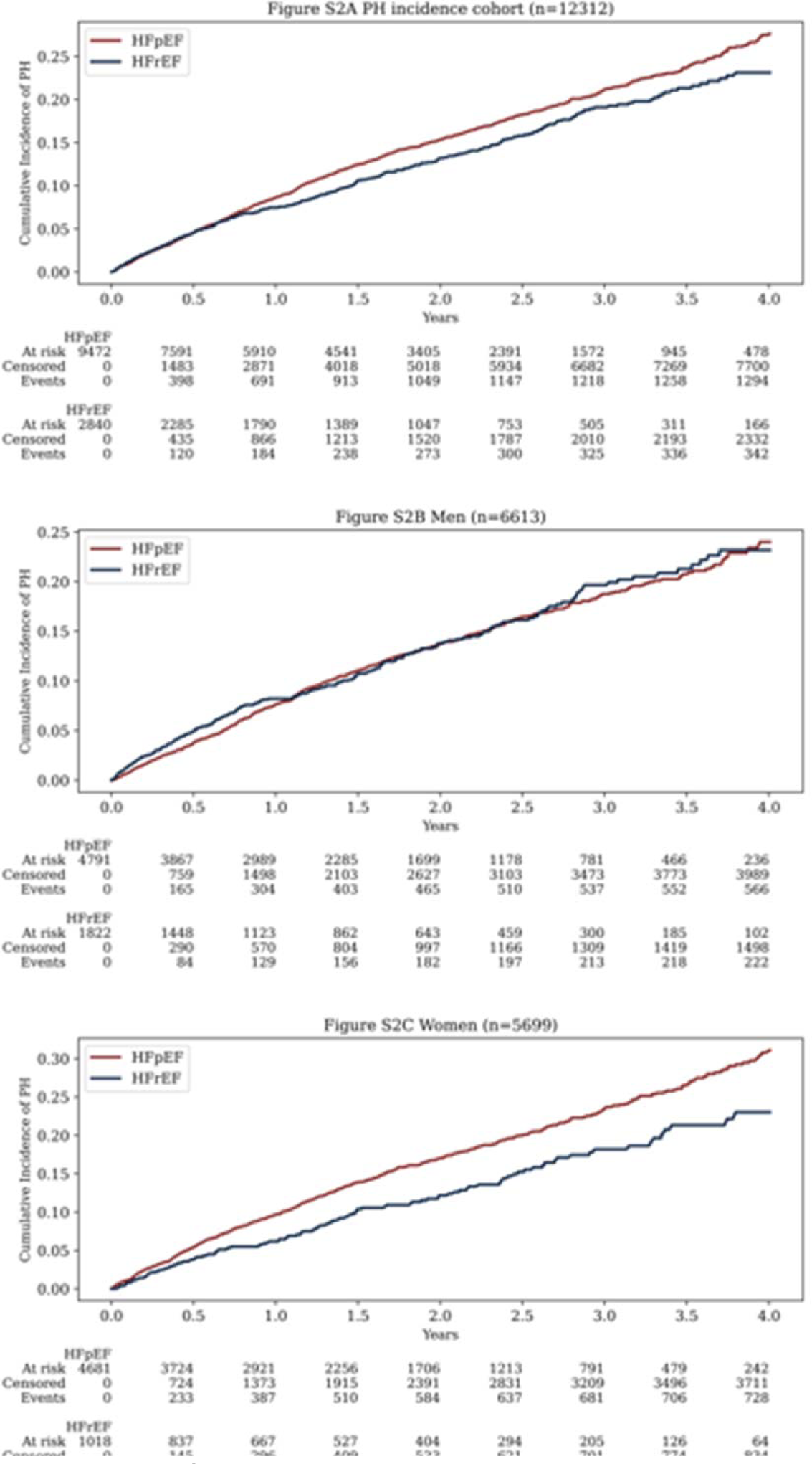
Cumulative Incidence of Group 2 pulmonary hypertension in HF-PH Association Cohort and for Men and Women.

**Supplemental Table 1:**
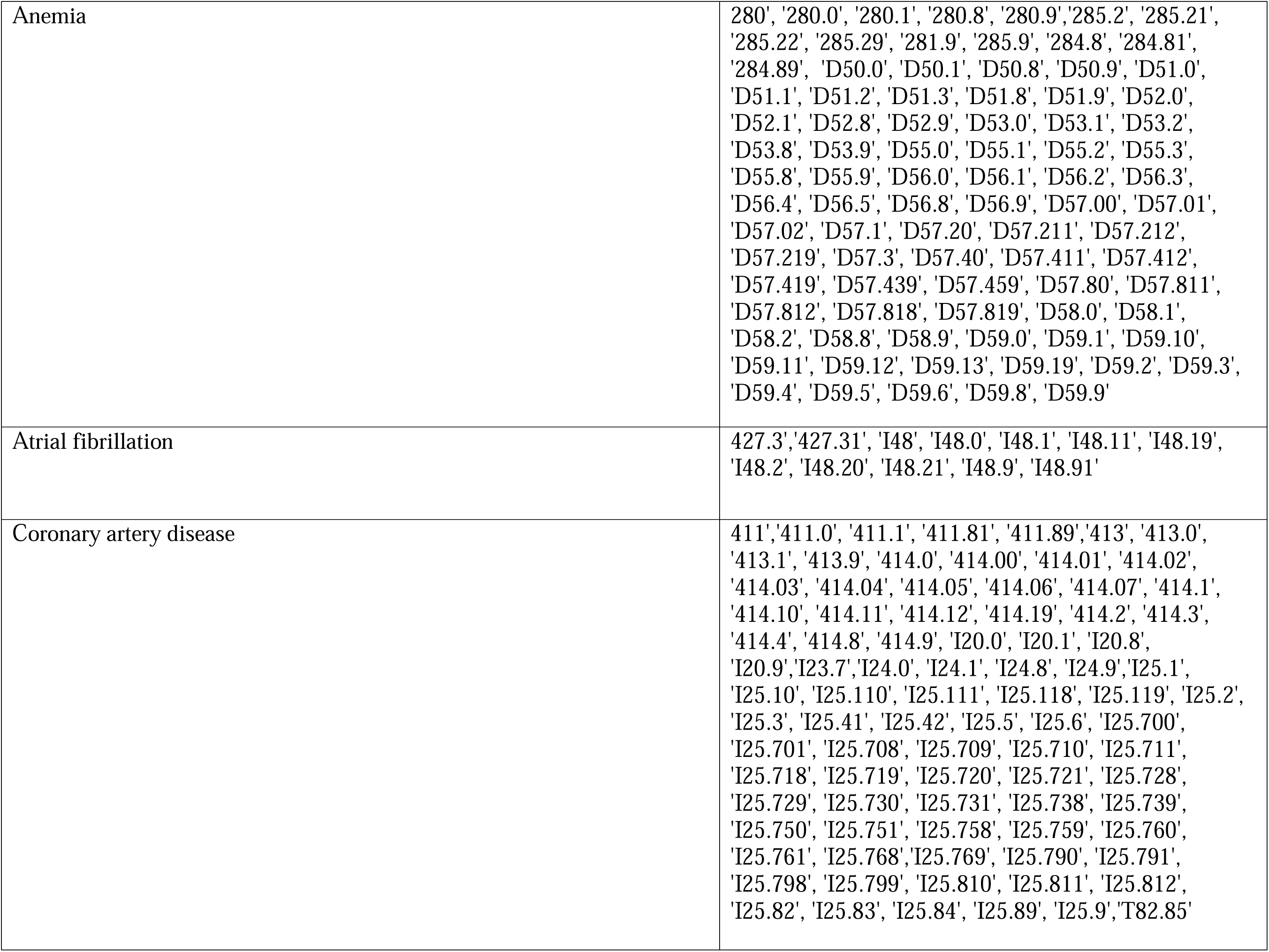
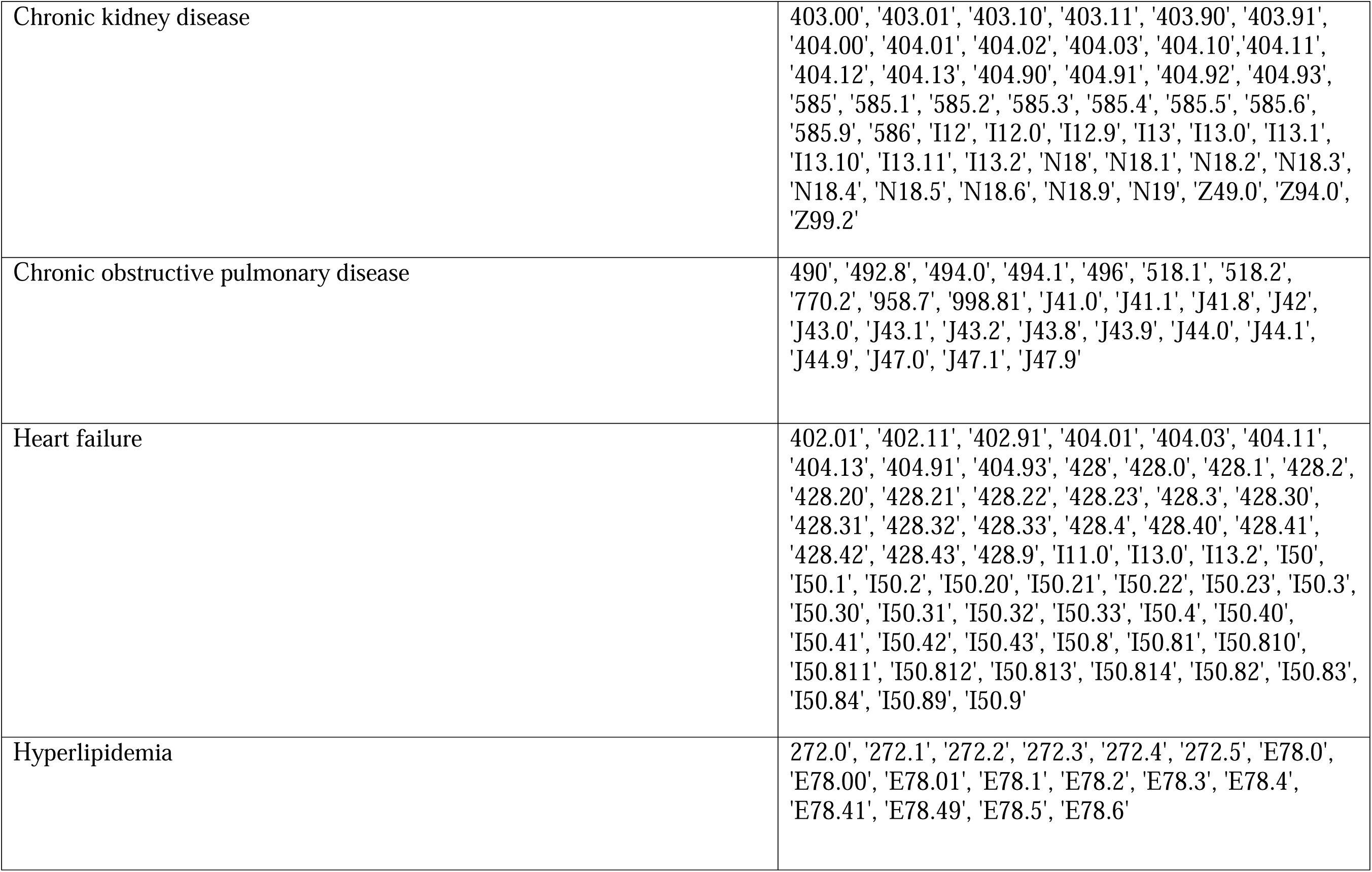
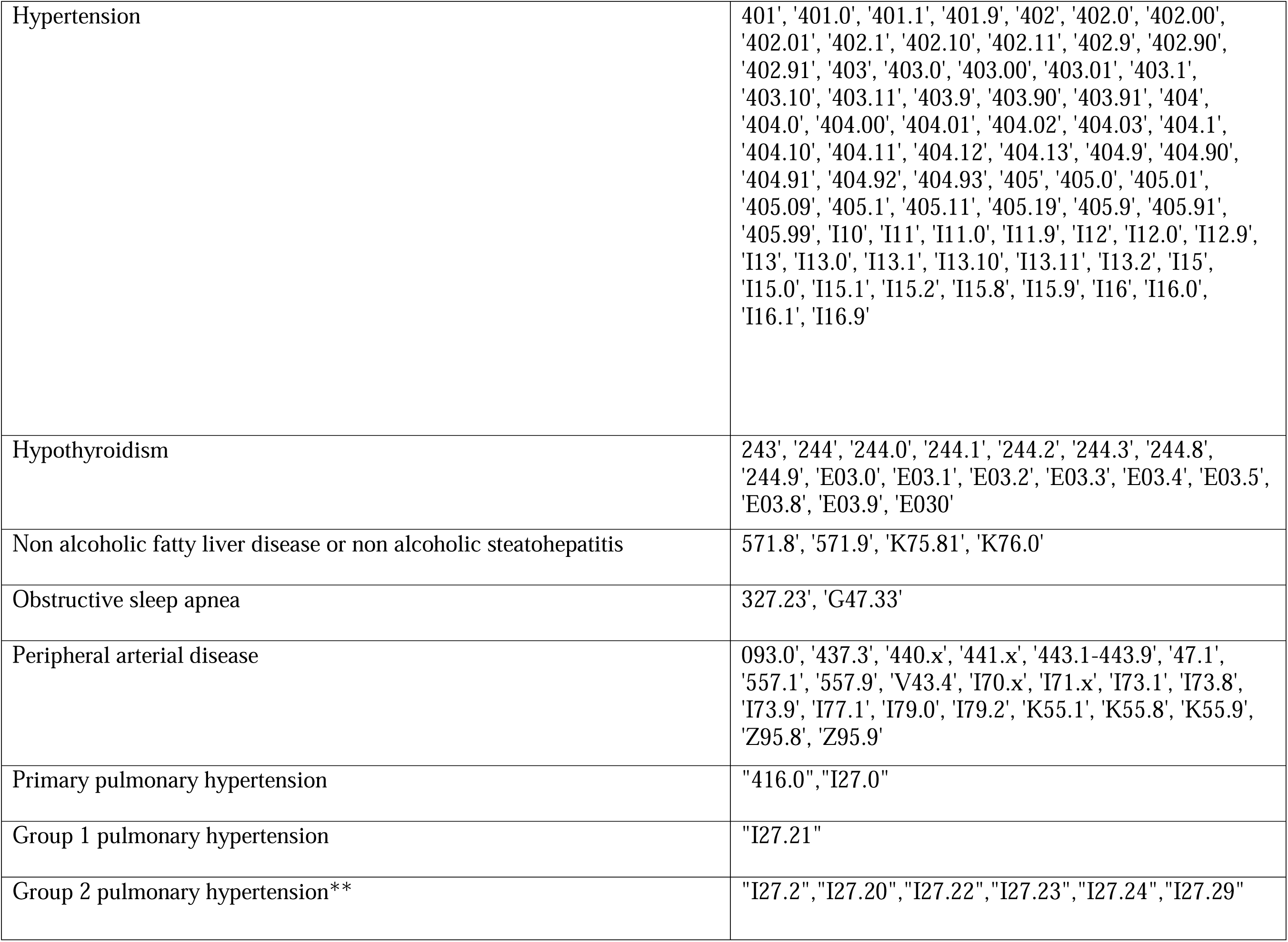
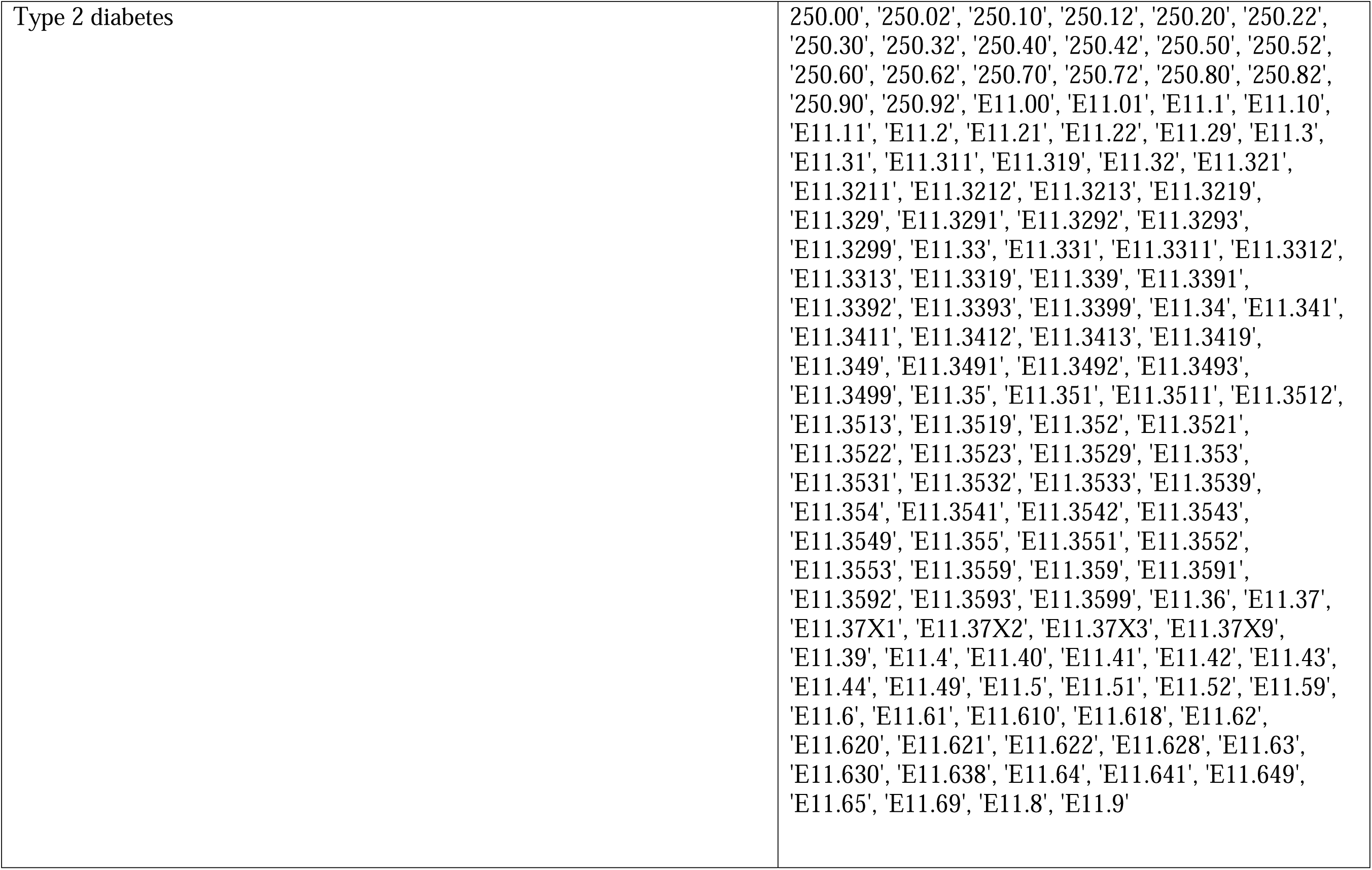
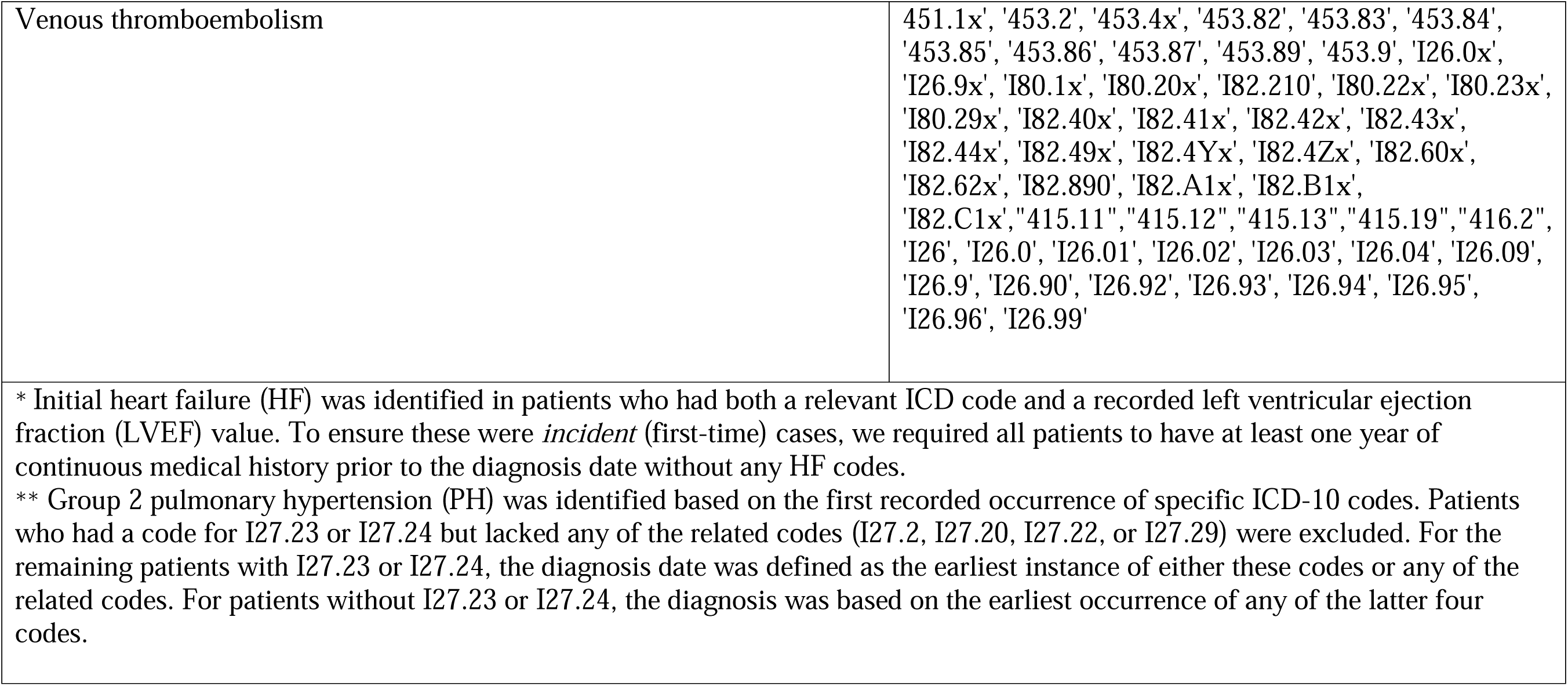
List of ICD-9 and ICD-10 Codes Used to Define Comorbidities and Outcomes.

**Supplemental Table 2:** Demographics and Characteristics of the PH Incidence Cohort.

|  |  | **Entire PH Incidence Cohort** | **No incident PH** | **Incident PH** | ***p* value** | **HFpEF** |
| --- | --- | --- | --- | --- | --- | --- |
| n |  | 17212 | 14191 | 2950 |  | 13083 |
| Age, mean (SD) |  | 72.4 (13.1) | 72.1 (13.0) | 74.0 (12.8) | <0.005 | 73.1 (12.8) |
| Gender, n (%) | Female | 7908 (45.9) | 6351 (44.8) | 1517 (51.4) | <0.005 | 6424 (49.1) |
|  | Male | 9304 (54.1) | 7840 (55.2) | 1433 (48.6) |  | 6659 (50.9) |
| Race, n (%) | White | 15638 (90.9) | 12957 (91.3) | 2617 (88.7) | <0.005 | 11971 (91.5) |
|  | Black | 1392 (8.1) | 1086 (7.7) | 299 (10.1) |  | 972 (7.4) |
|  | Other | 182 (1.1) | 148 (1.0) | 34 (1.2) |  | 140 (1.1) |
| BMI, mean (SD) |  | 30.9 (7.9) | 30.8 (7.8) | 31.2 (8.0) | 0.01 | 31.3 (8.1) |
| HF subtype, n (%) | HFpEF | 13083 (76.0) | 10778 (75.9) | 2237 (75.8) | 0.91 | N/A |
|  | HFrEF | 4129 (24.0) | 3413 (24.1) | 713 (24.2) |  |  |
| Anemia, n (%) |  | 6905 (40.1) | 5574 (39.3) | 1302 (44.1) | <0.005 | 5610 (42.9) |
| Atrial fibrillation, n (%) |  | 7224 (42.0) | 5838 (41.1) | 1365 (46.3) | <0.005 | 5586 (42.7) |
| Coronary arterial disease, n (%) |  | 11732 (68.2) | 9775 (68.9) | 1931 (65.5) | <0.005 | 8714 (66.6) |
| Chronic kidney disease, n (%) |  | 6004 (34.9) | 4817 (33.9) | 1167 (39.6) | <0.005 | 4701 (35.9) |
| Chronic obstructive pulmonary disease, n (%) |  | 7097 (41.2) | 5816 (41.0) | 1245 (42.2) | 0.23 | 5639 (43.1) |
| Hyperlipidemia, n (%) |  | 14586 (84.7) | 12080 (85.1) | 2462 (83.5) | 0.02 | 11251 (86.0) |
| Hypertension, n (%) |  | 16277 (94.6) | 13406 (94.5) | 2814 (95.4) | 0.05 | 12489 (95.5) |
| Hypothyroidism, n (%) |  | 4721 (27.4) | 3846 (27.1) | 866 (29.4) | 0.01 | 3795 (29.0) |
| Nonalcoholic fatty liver disease or nonalcoholic steatohepatitis, n (%) |  | 1433 (8.3) | 1211 (8.5) | 216 (7.3) | 0.03 | 1180 (9.0) |
| Obstructive sleep apnea, n (%) |  | 4221 (24.5) | 3500 (24.7) | 707 (24.0) | 0.44 | 3453 (26.4) |
| Peripheral arterial disease, n (%) |  | 5790 (33.6) | 4765 (33.6) | 1009 (34.2) | 0.53 | 4618 (35.3) |
| Type 2 diabetes, n (%) |  | 8207 (47.7) | 6674 (47.0) | 1512 (51.3) | <0.005 | 6325 (48.3) |
| Venous thromboembolism, n (%) |  | 1893 (11.0) | 1558 (11.0) | 314 (10.6) | 0.62 | 1532 (11.7) |

Values are presented as mean ± standard deviation or number (%); PH= pulmonary hypertension; N/A: not applicable

**Supplemental Table 3:** Cumulative Incidence (%) of Group 2 Pulmonary Hypertension After Heart Failure, Stratified by Sex Within Each HF Subtype.

| Time from HF | All | HFpEF |  | HFrEF |  |
| --- | --- | --- | --- | --- | --- |
|  |  | Women | Men | Women | Men |
| 30 days | 3.08 (2.82–3.33) | 3.30 (2.87–3.73) | 2.41 (2.04–2.77) | 3.80 (2.84–4.76) | 3.82 (3.11–4.54) |
| 90 days | 5.80 (5.46–6.14) | 6.28 (5.71–6.85) | 4.72 (4.21–5.22) | 7.01 (5.74–8.27) | 6.67 (5.75–7.59) |
| 180 days | 8.60 (8.19–9.00) | 9.21 (8.53–9.88) | 7.10 (6.49–7.70) | 10.89 (9.37–12.38) | 9.62 (8.53–10.69) |
| 1 year | 12.85 (12.37–13.33) | 14.17 (13.36–14.96) | 10.71 (9.97–11.44) | 14.38 (12.68–16.05) | 14.11 (12.84–15.37) |
| 2 years | 19.63 (19.05–20.21) | 21.83 (20.87–22.78) | 17.40 (16.47–18.33) | 20.14 (18.15–22.09) | 19.34 (17.86–20.80) |
| 3 years | 24.78 (24.11–25.45) | 27.40 (26.32–28.47) | 22.42 (21.31–23.51) | 24.45 (22.19–26.65) | 24.19 (22.47–25.88) |
| 4 years | 29.85 (29.03–30.67) | 33.42 (32.10–34.71) | 26.52 (25.16–27.86) | 29.87 (27.10–32.53) | 28.84 (26.75–30.87) |
| 4.5 years | 32.86 (31.88–33.83) | 37.40 (35.82–38.95) | 29.39 (27.71–31.02) | 31.37 (28.35–34.27) | 30.54 (28.23–32.77) |

The cumulative incidence rate of group 2 pulmonary hypertension in the entire PH incidence cohort after the index date.
HF: heart failure; HFrEF: heart failure with reduced ejection fraction; HFpEF: heart failure with preserved ejection fraction

